# A Data-Driven Framework for Generating Population-Linked Case Vignettes from Nationwide Triage Data

**DOI:** 10.64898/2026.06.08.26354886

**Authors:** Anja Seidel, Edgar Steiger, Josephine Schuster, Lars Eric Kroll

**Affiliations:** Department of IT and Data Science, Central Research Institute of Ambulatory Health Care (Zi), Salzufer 8, 10587, Berlin, Germany; Department of Medicine, Central Research Institute of Ambulatory Health Care (Zi), Salzufer 8, 10587, Berlin, Germany

**Keywords:** Data Science, Medical Informatics, Clinical Decision Support Systems, Triage, Machine Learning, Clustering Algorithms, Natural Language Processing, Large Language Models

## Abstract

**Background:** Digital decision-support tools such as triage systems and symptom checkers support millions of health-related decisions each year. Their quality and safety are commonly evaluated using textual patient cases, known as case vignettes. However, existing vignette sets written by medical experts cover only a limited spectrum of real-world patient presentations and lack population weights, which would allow extrapolating evaluation results to the underlying patient population.

**Objective:** This study aims to develop a data-driven framework for automatically generating a human-manageable set of case vignettes from nationwide triage data that captures broad presentation diversity and links each vignette to a quantitative weight reflecting the number of underlying patient assessments.

**Methods:** From 3.2 million triage assessments conducted over one year using structured triage software in the German medical on-call service (telephone triage and online self-triage) and at the joint contact points of the outpatient emergency care service and hospital emergency departments, we randomly sampled 50,000 cases. Triage questionnaires were converted into semantic embeddings using a German Sentence Transformer Model and grouped by agglomerative clustering. For clusters containing sufficient assessments, we generated one representative assessment using a two-phase simulated-annealing optimization. The optimization minimized the distance to the cluster centroid while maximizing the number of answered triage questions, aiming for high representativeness and information content. Each representative assessment was assigned the size of its source cluster as its sample-based weight. A similarity-based sensitivity analysis was performed to examine whether these weights were preserved in the full 1-year population. Finally, the question-answer pairs of the representative assessments were converted into structured textual case vignettes using controlled prompting of a large language model.

**Results:** The cluster analysis yielded 514 included clusters covering 96.8% of the sampled 50,000 assessments. The generated representatives showed strong agreement with the majority treatment-urgency recommendation of their source cluster (Spearman’s ρ=0.78, p<0.001) and contained on average 4.3 more answered triage questions than the original assessments within their clusters. When weighted by cluster size, the representatives approximated the sample distributions of treatment urgency, demographics, and symptoms, although some systematic deviations remained, most notably an overrepresentation of female cases (+13.5%), patients aged 14-49 years (+8.0%), and the urgency category “As soon as possible” (+6.6%). Of 121 recorded symptoms, 101 (83.5%) were covered by the representatives; the rest each occurred in <0.5% of the sample. In a sensitivity analysis, cluster-based vignette weights were strongly correlated with similarity-based population weights (Spearman’s ρ=0.77, p<0.001), and 90.1% of assessments in the full 1-year population were matched to at least one vignette.

**Conclusions:** We present a data-driven framework for deriving a manageable set of population-weighted case vignettes from nationwide triage data. The resulting vignettes captured broad presentation diversity, approximated key sample characteristics, and provided an explicit quantitative link to the number of underlying patient assessments. After medical expert review and refinement, the vignettes may support more population-aware evaluation and quality assurance of digital decision-support tools.

## Introduction

### Background

Digital decision-support tools, such as triage systems and symptom checkers, are increasingly used by both healthcare professionals and medical laypeople [1–3]. Beyond stand-alone consumer applications, triage systems also operate within routine care pathways, including nationwide services, where they support treatment decisions in emergency medical services and on-call medical services. In Germany, for example, the on-call medical service is supported by a software-as-a service triage solution that conducted around 3.1 million telephone triage and online self-triage assessments in 2024 [3] and more than 3.4 million in 2025 [3].

The widespread use of these tools and the large number of patients affected underscore the importance of ongoing quality assurance. In addition, given that an increasing number of alternative products are available which, despite conforming to medical device regulations, appear to achieve different and partly conflicting results (e.g. [4,5]), national agencies need to make informed decisions concerning the choice of triage software. In triage settings, accurate identification of urgent cases is essential to ensure timely referral to appropriate medical care, whereas less-urgent cases should be de-escalated accordingly. Errors in either direction can have major consequences: under-triage, in which a serious case is classified as not urgent enough, can lead to health- or even life-threatening delays, whereas over-triage, in which patients are assessed as too urgent, unnecessarily consumes limited health care resources, and may delay treatment of truly urgent patients.

Because many triage tools and symptom checkers are proprietary rather than open source, a common method to evaluate such tools is through medical case vignettes - a textual description of specific patient cases. These vignettes are entered into triage systems or symptom checkers, and the resulting recommendations are compared with those made by medical professionals. In case of discrepancies, correcting adjustments can be made to the system under evaluation. Using this approach, the comparison between systems and their further development depends heavily on the set of vignettes used.

### Prior Work

Previous studies have often relied on case vignettes authored by medical professionals and based on real cases or adapted from teaching materials and the medical literature (e.g., [6–10]). Such expert-written vignettes can provide high medical quality, but their limited number usually constrains the range of patient presentations represented. As a result, they may not adequately reflect the diversity of real-world presentations encountered by triage systems or symptom checkers.

To increase realism and breadth, other studies have sampled real patient cases and converted them into vignettes (e.g., [11–13]). These approaches are more likely to capture broader clinical variation, but explicit evaluation of how well the resulting vignette sets reflect population-level characteristics, such as demographic distributions or symptom frequencies, are rare [12]. In addition, random sampling may omit rare but medically critical conditions while over-sampling common, similar cases. Other work has leveraged entire electronic health record datasets available for a defined patient group [14], but such approaches are feasible only for limited cohort sizes.

More recently, large language models (LLMs) have been used to generate medical vignettes (e.g., [15–18]). While some of these studies focused on predefined disease areas (e.g., [16,18]), none aimed to capture or analyze population-wide characteristics of patient presentations. Moreover, to our knowledge, prior vignette studies have generally not quantified the representativeness of individual vignettes in terms of how many real-world patients each vignette corresponds to within the underlying population. Such information would be highly valuable, as it could support extrapolation of evaluation results, such as misclassification rates, from the vignette set to the expected number of affected patients in the population. Thus, an important gap remains: there is currently no established framework for deriving a human-manageable vignette set from large-scale patient cohorts while preserving broad presentation diversity and linking individual vignettes to explicit quantitative weights.

### Scope and Objectives

In this study, we used nationwide triage data comprising all patient contacts with the on-call medical service (116117 phone line or website) in Germany. Patients contacting this service typically seek care for medical problems they perceived as urgent but not immediately life-threatening. The resulting triage recommendations, nevertheless, can span a broad range, from emergency referral to non-urgent telemedical consultation.

The aim of this study was to develop a data-driven framework for generating a set of medical vignettes that would

i. capture broad characteristics of most 116117 contacts in terms of population characteristics such as demographics and symptom patterns,
ii. include an explicit weight reflecting the number of underlying patient assessments represented by each vignette,
iii. ensure explicit consideration of rare emergency cases given the serious potential consequences of their misclassification, and
iv. remain manageable in size to enable subsequent medical-expert review and quality control.

To the best of our knowledge, no prior study has described a framework for deriving a human-manageable set of vignettes from large-scale real-world data that retains near-complete sample coverage and links each vignette to an explicit sample-based weight.

To achieve these objectives, the questionnaires from a large sample of real triage assessments were semantically clustered, and one representative assessment was generated for eligible clusters. Because each representative assessment originated from a defined cluster, the number of underlying assessments, and thus its weight relative to the sample population, was known. Subsequently, each representative assessment was converted into a structured textual vignette using controlled prompting of a recent large language model (LLM).

## Methods

### Framework Overview

Our proposed framework for generating a condensed, reviewable set of population-linked case vignettes from nationwide triage data is shown in Figure 1. First, triage questionnaires from a random sample of 50,000 real-world assessments collected over one year were transformed into semantic embeddings and grouped by agglomerative clustering to identify groups of similar patient presentations. For clusters containing a sufficient number of assessments, one representative assessment was generated to serve as the basis for a single vignette. Each representative assessment was intended to capture the central characteristics of its cluster while also resembling a realistic patient case that responds to many triage system questions. To balance these two aims - high representativeness and high information content - we implemented a two-phase optimization procedure based on simulated annealing. Because each representative assessment originated from a defined cluster, the number of underlying assessments and thus its weight relative to the sample population, was known. This enabled comparison of the distributions of demographics, symptoms, and triage recommendations between the representative cases and the full sample. To assess whether the clustering-based weights are transferable to the full population, we additionally performed a similarity-based sensitivity analysis in the full 1-year assessment dataset. Finally, to make the results easily interpretable by medical professionals, every representative assessment was converted into a structured textual vignette using controlled prompting of a large language model (LLM).

**Figure 1:**
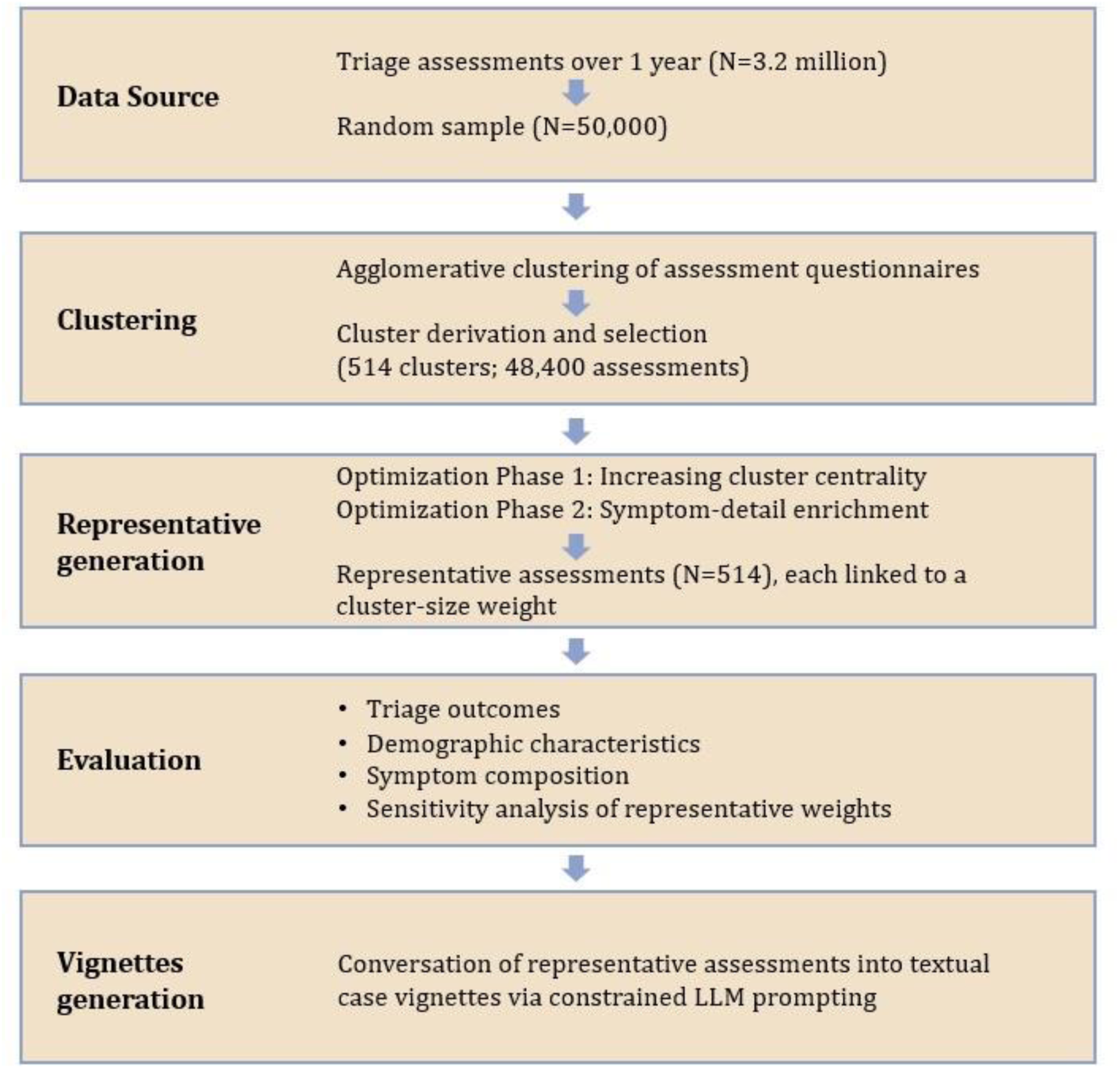
Overview of the study framework for generating population-linked case vignettes from nationwide triage data.

### Data Source

The data for this study originated from patient contacts with the German medical on-call service (116117) and from joint contact points of the outpatient emergency care service and hospital emergency departments (“Gemeinsamer Tresen” in German). Patients entered the triage pathway either by calling the hotline, where information was collected during the call; by completing an online self-assessment questionnaire; or by presenting in person at a joint contact point, where an on-site assessment was conducted. In all settings, versions of the same structured triage software based on clinical guidelines and published evidence were used. The system applies a standardized question flow, stores patient-reported information as an assessment, and generates a triage recommendation specifying the urgency of treatment and point of care. To account for seasonal effects, we considered all assessments recorded over a period of one year (1 November 2023 until 31 October 2024). From 3.2 million available assessments, we randomly selected a subset of 50,000. Each assessment contained the following information: date of assessment, patient’s age, gender, pregnancy status (if applicable), details about reported medical symptoms, responses to general health questions (e.g. previous hospital visits, number of medications, pre-existing conditions) and the triage outcomes time to treat (TTT) and point of care (POC). TTT comprised the categories “Emergency”, “As soon as possible”, “Within 24 hours”, “Not required within 24 hours” and “Unclear or N/A”; POC comprised “Emergency ambulance”, “Emergency room”, “Physician”, “Teleconsultation” and “Unclear or N/A”. Descriptive characteristics of the full 1-year dataset and the random sample are reported in Table 1 in the Results.

**Table 1.** Demographic and triage-system characteristics of the full 1-year assessment dataset and the random sample of 50,000 assessments.

| Characteristics | 1-year<br>(N=3,166,727) | Sample<br>(N=50,000) |
| --- | --- | --- |
| **Demographics** |  |  |
| **Gender, n (%)** |  |  |
| Male | 1,310,551 (41.4) | 21,120 (42.2) |
| Female | 1,820,554 (57.5) | 28,692 (57.4) |
| N/A | 35,622 (1.1) | 188 (0.4) |
| **Age, n (%)** |  |  |
| 1-4 Weeks | 4,735 (0.1) | 82 (0.2) |
| 5-8 Weeks | 5,660 (0.2) | 92 (0.2) |
| 3-12 Months | 55,613 (1.8) | 873 (1.7) |
| 1-3 Years | 157,942 (5.0) | 2,414 (4.8) |
| 4-8 Years | 139,096 (4.4) | 2,190 (4.4) |
| 9-13 Years | 74,190 (2.3) | 1,114 (2.2) |
| 14-49 Years | 1,191,704 (37.6) | 19,790 (39.6) |
| 50-65 Years | 471,641 (14.9) | 7,375 (14.8) |
| 66-80 Years | 477,452 (15.1) | 7,372 (14.7) |
| >80 Years | 548,984 (17.3) | 8,448 (16.9) |
| N/A | 39,710 (1.3) | 250 (0.5) |
| **Triage Outcomes** |  |  |
| **TTT, n (%)** |  |  |
| Emergency | 224,443 (7.1) | 3,745 (7.5) |
| As soon as possible | 1,379,244 (43.6) | 21,966 (43.9) |
| Within 24 hours | 959,591 (30.3) | 14,929 (29.9) |
| Not required within 24 hours | 444,232 (14.0) | 7,010 (14.0) |
| Unclear or N/A | 159,217 (5.0) | 2,350 (4.7) |
| **POC, n (%)** |  |  |
| Emergency ambulance | 158,267 (5.0) | 2,696 (5.4) |
| Emergency room | 809,130 (25.6) | 13,324 (26.6) |
| Physician | 1,502,027 (47.4) | 23,373 (46.7) |
| Teleconsultation | 531,630 (16.8) | 8,153 (16.3) |
| Unclear or N/A | 165,673 (5.2) | 2,454 (4.9) |
| **Triage Questionnaires** |  |  |
| **Symptoms** |  |  |
| n available | 121 | 121 |
| n selected<br>(median, mean, SD) | 1, 1.7, 1.3 | 1, 1.7, 1.3 |
| **Questions** |  |  |
| n answered (median,<br>mean, SD) | 15, 16.0, 9.5 | 15, 16.1, 9.5 |

### Ethical Considerations

The data used in this study consist of pseudonymized individual-level social data that were lawfully collected and stored in accordance with Section 285(3) in conjunction with Section 285(1) No. 6 of the German Social Code, Book V (SGB V), and may be processed for quality assurance purposes. The data are owned by the Associations of Statutory Health Insurance Physicians (ASHIPs). On the basis of contractual data processing agreements pursuant to Section 80 of German Social Code, Book X (SGB X) in conjunction with Article 28 of the General Data Protection Regulation (GDPR), the Central Research Institute of Ambulatory Health Care in Germany (Zi) processes these data in pseudonymized form for quality assurance. Data transfer and processing were reviewed for compliance with applicable data protection regulations. Access to the data was restricted to authorized personnel.

### Clustering of Triage Assessments

To identify groups of assessments with similar patient-reported presentations, we performed a cluster analysis. Clustering was based exclusively on the information actively entered into the triage system, namely the questions and their corresponding answers, while triage system generated outcomes (TTT and POC) were not considered. This separation was intended to ensure that the clustering reflects the semantic similarities in patient-reported information and allows the TTT to be used subsequently as an external criterion for evaluating cluster quality.

Because the ideal number of clusters is unknown, we applied agglomerative clustering [19], which builds a dendrogram and does not require specifying the number of clusters in advance. Since agglomerative clustering scales quadratically with the number of assessments in both memory and runtime, applying it to the 1-year population of more than 3 million assessments was computationally infeasible. We therefore clustered the random sample of 50,000 assessments as a pragmatic compromise between population coverage and computational resources.

For agglomerative clustering, we systematically evaluated several design choices: the representation of the question-answer pairs (one-hot encoding, one-hot encoding followed by principal component analysis (PCA) [20], and semantic embeddings generated with the Sentence Transformer model “German Semantic STS V2” [21]), the linkage method (“average”, “complete”, “ward”) and the distance metric (“euclidean”, “cosine”, “jaccard”). Only eligible combinations were evaluated.

To compare the resulting configurations, we assessed TTT homogeneity over different numbers of clusters (i.e., different dendrogram cuts, see Figure A.1 in Multimedia Appendix 1). TTT homogeneity ranges from 0 to 1, with higher values indicating that assessments within clusters more often share the same TTT. Because clinically similar presentations would be expected to yield similar urgency recommendations, we used TTT homogeneity as an external proxy for one clinically relevant aspect of clustering quality. For each configuration, we computed the TTT homogeneity score across a range of 1 to 5,000 clusters. Note that a higher number of clusters results in grouping of more similar assessments and therefore higher TTT homogeneity. We then selected the clustering configuration that achieved a TTT-homogeneity score of 0.5 with the smallest number of clusters. Although higher homogeneity would be desirable in principle, this would result in substantially more clusters (with considerably fewer assessments per cluster) and consequently a much higher number of vignettes to be generated (e.g., a homogeneity of 0.6 would result in >2.500 clusters, see Figure A.1 in Multimedia Appendix 1). A homogeneity threshold of 0.5 was therefore chosen as a trade-off between medical similarity of assessments within clusters and the number and size of clusters obtained.

Under this selection criterion, the best clustering performance was achieved using semantic embeddings from the Sentence Transformer model “German Semantic STS V2” [21] combined with Ward linkage and Euclidean distance. “German Semantic STS V2” is specifically trained to assess semantic similarity between German texts. Each assessment was represented as a text document by concatenating all answered question-answer pairs and prefixing symptom-specific ones with the symptom name. The resulting text documents were encoded into 1024-dimensional embeddings using “German Semantic STS V2” and then clustered using the cluster.hierarchy.linkage function from the SciPy package version 1.15.2 [22] with parameter method=”ward” and metric=”euclidean”.

For representative assessment generation, we retained clusters containing at least 30 assessments. This minimum cluster size was chosen as a pragmatic compromise between retaining broad sample coverage and ensuring that each retained cluster contained a meaningful number of assessments to build a representative. Higher minimum cluster-size thresholds would have reduced sample coverage substantially (see Supplementary Table A.1 in Multimedia Appendix 1). Because emergency cases are of particular medical relevance but occurred less frequently in our sample, we additionally included clusters exclusively comprising emergency assessments (POC of all assessments in [“Emergency ambulance”, “Emergency room”]).

### Generation of Representative Assessments

After identifying clusters of similar assessments, we aimed to construct one representative assessment per cluster. This representative case was intended to (1) reflect the central characteristics of the cluster and to (2) contain as much information as possible, i.e. include as many answered triage-system questions as possible - without losing representativeness. Emphasizing the second criterion ensured that the resulting assessment not only captured dominant characteristics of the cluster but also resembled a plausible case in which many questions of the triage system could be answered.

#### Phase 1: Heuristic Search for Representative Cluster Assessments

For each cluster, we aimed to build one artificial representative assessment that best captures the cluster’s key characteristics. First, the cluster centroid was computed as the mean of all 1024-dimensional sentence-embedding vectors of the assessments within the cluster. Next, the Euclidean distance between each assessment’s embedding in the cluster and the centroid was calculated, and the assessment with the smallest distance to the centroid was selected as the initial representative assessment. Subsequently, the frequency distribution of all question-answer combinations observed within the cluster, i.e., the proportion of assessments in which each combination occurred, was determined. For each question occurring in the cluster, we additionally introduced an explicit “unanswered” state with probability corresponding to the proportion of cluster assessments in which that question has not been answered. Based on this frequency distribution, the initial assessment was iteratively refined using simulated annealing, a stochastic optimization algorithm that searches for a global minimum by occasionally accepting worse intermediate solutions to escape local minima. In each iteration, one question-answer pair was sampled according to the cluster-specific frequency distribution, and the representative assessment was edited to include this sampled answer. Depending on the question type and previous answers, this edit could replace the currently selected answer to a question, add an answer to a previously unanswered question, or remove an answer by setting the respective question to the “unanswered” state. Additional rule-based constraints were enforced to preserve questionnaire consistency. When a symptom was removed by setting a symptom-presence question to “no” or “unanswered”, all associated symptom-specific detail questions were removed as well (set to “unanswered”). Conversely, when a symptom-specific detail question was answered, the corresponding symptom-presence question was automatically set to “yes” if not already present. Symptom combinations were restricted to those permitted by the age- and gender-specific symptom structure of the triage system, and changes in age or gender triggered re-evaluation of symptom validity. Pregnancy-related questions were only allowed for female patients in the reproductive age range. Finally, dependent child questions were removed whenever their required parent question-answer condition was no longer fulfilled.

After each updating step, the embedding of the modified assessment was built, and its distance to the centroid was re-computed. Simulated annealing was run for up to 10.000 iterations, starting with an initial temperature of 1.000, a minimum temperature of 0.0001, and cooling factor alpha of 0.95. The generated assessment achieving the smallest distance to the centroid over all iterations was selected as the representative assessment for that cluster. The resulting representative assessments per cluster were then enriched into more detailed assessments, as described in the following section.

#### Phase 2: Enrichment of Representative Cluster Assessments

The representative assessments obtained in Phase 1 captured key features of each cluster but can lack details regarding the reported symptoms. Therefore, we performed an additional enrichment step aimed at increasing the information density and realism of each representative assessment while preserving the underlying symptom patterns identified in Phase 1. In this second phase, the symptom set of the Phase 1 representative assessment was kept fixed, and only symptom-specific detail questions belonging to already selected symptoms were allowed to change. For each selected symptom, a relative importance weight was calculated according to Formula 1 by removing all information related to that symptom from the representative assessment, recomputing the assessment’s embedding and the distance to the cluster centroid. Removing important symptoms increased the distance to the centroid more than removing less important ones.

After obtaining the symptom weights, we re-ran simulated annealing with the same parameters as in Phase 1, starting from the Phase 1 representative assessment. In each iteration, one answer was sampled from the empirical cluster-specific distribution of symptom-detail answers for the already selected symptoms, again including an explicit “unanswered” state. The generated assessments were evaluated using an adapted cost function (Formula 2) that balanced proximity to the cluster centroid against the number of answered symptom-related questions, weighted by symptom importance.

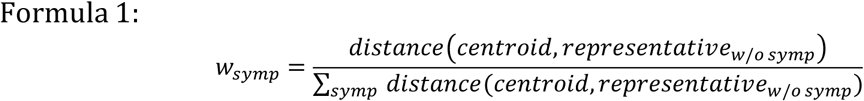

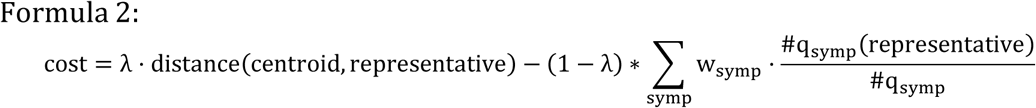

The assessment that achieved the lowest cost value was retained as the final representative assessment for that cluster. #q_symp(representative) denotes the number of symptom-specific detail questions answered for symptom “symp” in the current representative assessment, and #q_symp denotes the number of such questions that were observed with an answer in the source cluster for that symptom. The weighting factor λ in [0,1] controls the trade-off between minimizing the distance to the centroid and maximizing the number of answered symptom-related questions. We selected λ=0.25 to favor more complete triage questionnaires while maintaining proximity to the cluster centroid. In a sensitivity analysis comparing λ=0.25 and λ=0.50, differences in centroid distance were small, whereas λ=0.25 yielded slightly richer representative assessments (see Supplementary Table A.2 in Multimedia Appendix 1).

### Evaluation of Representative Assessments

#### Agreement with Cluster-Level Triage Urgency

Each representative assessment originated from an actual case within its cluster and was adjusted using the frequency distributions of the respective question-answer combinations, ensuring that all assessment characteristics occurred in the cluster.

Nevertheless, different answer combinations can correspond to varying symptom patterns and severity levels. Therefore, a reliable representative assessment should not only capture dominant symptom patterns of its cluster, but also reflect the cluster’s majority triage recommendations, i.e., the dominant treatment urgency. To evaluate this, each representative assessment was processed through the same triage system used for the original assessments by automatically submitting the corresponding question-answer pairs through the system’s application programming interface (API). The resulting triage outcomes (e.g., TTT) of each representative assessment were recorded and compared with the TTT distribution of the originating cluster.

#### Symptom Coverage and Symptom Category Mapping

The triage system recorded information on 121 distinct symptoms. To evaluate how well the representative assessments covered the symptom spectrum observed in the original data, these symptoms were grouped into 13 broader symptom categories, loosely following the structure of the “10th revision of the International Classification of Diseases – German Modification” (ICD-10-GM) [23] and reviewed by a medical expert. Each symptom was assigned to one most suitable category. No suitable mapping could be identified for the general symptoms “Fatigue”, “Feeling unwell” and “Crying baby/toddler”. The complete mapping is provided in Supplementary Table A.3 in Multimedia Appendix 2.

#### Sensitivity Analysis of Vignette Weights

In our primary approach, each representative assessment and its resulting vignette were directly linked to a cluster of real-world assessments of known size, thereby providing a native cluster-based weight within the 50,000 assessment sample.

However, this linkage may become less directly interpretable after medical expert review of the vignettes or under changing patient populations and is not transferable to external vignette sets. Therefore, we conducted a similarity-based sensitivity analysis that estimated vignette weights in the 1-year population of approximately 3.2 million assessments without relying on the original cluster-based weighting.

For this purpose, each assessment from the generated vignette set and each assessment from the 1-year population was represented as a text document by concatenating the question-answer pairs and prefixing symptom-specific entries with the corresponding symptom name, analogous to the procedure used in the cluster analysis. These documents were embedded into a 1024-dimensional vector using the “German Semantic STS V2” Sentence Transformer Model. We then computed the pairwise cosine similarity between each vignette assessment and all assessments in the 1-year population. To determine an appropriate similarity threshold, we performed a calibration analysis based on symptom overlap. For similarity thresholds between 0.90 and 1.00, we first identified all vignette-population pairs exceeding the respective threshold and then calculated the mean Jaccard index between their sets of recorded symptoms, defined over the 121 symptom labels defined in the triage system. Under the assumption that recorded symptoms of semantically similar assessments should at least moderately overlap, we selected the threshold at which the mean Jaccard index reached a value of 0.5, resulting in a similarity threshold of 0.955.

Using this approach, each population assessment could be matched to multiple vignettes from the same set. To avoid double counting, each matched population assessment was assigned a fractional weight inversely proportional to the number of similar matched vignettes (Formula 3). The similarity-based weight per vignette was then defined as the sum of the fractional weights of all similarly matched population assessments (Formula 4).

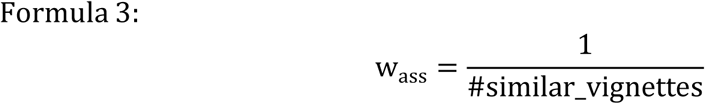

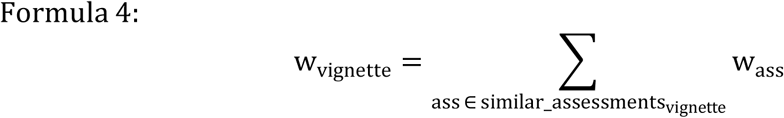

By construction, the sum of the similarity-based weights per vignette equals the total number of population assessments matched to at least one vignette above the selected similarity threshold. These similarity-based weights provide an alternative population-level estimate of vignette weight and were compared with the original cluster-based weights using Spearman rank correlation.

### Conversion of Representative Assessments into Textual Case Vignettes

After identifying one representative assessment per cluster, we aimed to formulate a specific patient vignette for each case using an LLM. We used GPT-4.1 (OpenAI, Generative Pre-Trained Transformers) and prompted it in German (because the triage questions were originally in German and we wanted to avoid language mixing) to generate a case vignette based on the question-answer pairs of each representative assessment. The model was instructed to:

- Use *only* the information contained in the given questions and answers and avoid adding assumptions or external knowledge (to prevent hallucinations).
- Include *all* answered questions in the vignette, regardless of apparent relevance (to prevent information loss).
- Structure the vignette in three sections: 1. Patient information (age, gender, pregnancy, if applicable), 2. Medical history (structured by symptoms or symptom complexes), 3. Other medically relevant information (e.g., risk factors, medications, pre-existing conditions, lifestyle)
- Use a medically precise, factual style of language.
- Select a specific value for interval expressions by sampling from a uniform distribution for closed intervals (e.g., age between 9 and 13 years, pain intensity between 4 and 7), and from an asymmetric distribution for open intervals (e.g., more than 3 days, older than 80 years, body temperature > 30°C, blood pressure < 90mmHg) assigning higher probabilities closer to values near the stated boundary.

The final instruction, selecting precise values within intervals, was necessary because the representative assessments were derived from triage data, and the triage system often records information in broad categories rather than exact values (for instance, duration of symptoms, age, pain intensity, or vital signs). To simulate realistic patient cases, these categorical ranges had to be converted into specific values. While the LLM handled most interval expressions correctly, it occasionally retained interval expressions for vital signs (e.g., blood pressure between 120 and 140 mmHg). Therefore, we added a preprocessing step that algorithmically assigned precise numerical values for vital-sign-related questions. All triage questions capturing vital signs were identified through algorithmic analysis of their possible answers, and each affected representative assessment was assigned a random value within the corresponding range. Closed intervals were sampled from a uniform distribution, whereas open intervals were drawn from an exponential distribution within physiologically plausible limits for human vital signs.

The complete prompt is provided in Supplementary Methods A.1 in Multimedia Appendix 3 for transparency and reproducibility.

## Results

### Study Sample

From the full 1-year dataset of triage assessments recorded by the German medical on-call service, we randomly sampled 50,000 assessments for clustering and representative assessment generation. As shown in Table 1, the random sample closely resembled the full 1-year dataset with respect to demographic characteristics, triage recommendations, and triage questionnaires.

### Clustering Results

We evaluated several approaches to cluster the medical assessments of the sample, each represented by a set of question-answer pairs recorded in the triage system. Clustering quality was evaluated using TTT homogeneity as a proxy, because assessments with similar medical patterns should yield similar recommendations for treatment urgency. In the following, we present the outcomes of the best-performing configuration, defined as the one that achieved a TTT homogeneity of 0.5 with the fewest clusters. The performance of all tested clustering configurations is provided in Multimedia Appendix 1, Supplementary Figure A.1.

#### Dendrogram Characteristics

The best-performing clustering configuration was agglomerative clustering using semantic embeddings from the Sentence Transformer model “German Semantic STS V2”, combined with Ward linkage and Euclidean distance. The resulting dendrogram is shown in Figure 2. We analyzed the main branches (indicated by the dashed line) with respect to the distribution of reported symptoms and potentially urgent/emergency complaints recorded during the pre-triage phase (prefixed with “PT”) in the underlying assessments. The first two branches contain discontinued or incomplete assessments without triage recommendation or recorded symptoms (Branch 1 - left unbranched child of the root: number of assessments (n_ass): 487, Branch 2: n_ass: 1,899). The remaining branches correspond to distinct thematic topics:

- Branch 3: Musculoskeletal and cardiovascular complaints, led by lumbar back pain, leg problems and dizziness (n_ass: 10,100)
- Branch 4: Digestive-tract symptoms, led by vomiting/nausea, stomach pain, urinary tract complaints and diarrhea (n_ass: 9,637)
- Branch 5: Eye complaints, led by eye redness, eye pain and vision defect (n_ass: 3,035)
- Branch 6: Common cold-like symptoms, led by fever, cough, influenza infection (n_ass: 11,610)
- Branch 7: Comprises a right sub-branch with skin problems and injuries, led by rash, wound/skin injury, accident/trauma and itching, and a left sub-branch with mental-health issues, led by depressive feeling and anxiety (n_ass: 9,434)
- Branch 8: Acute and potential emergency conditions, many identified during pre-triage, such as cardiovascular, respiratory and neurological complaints (n_ass: 3,798)

**Figure 2:**
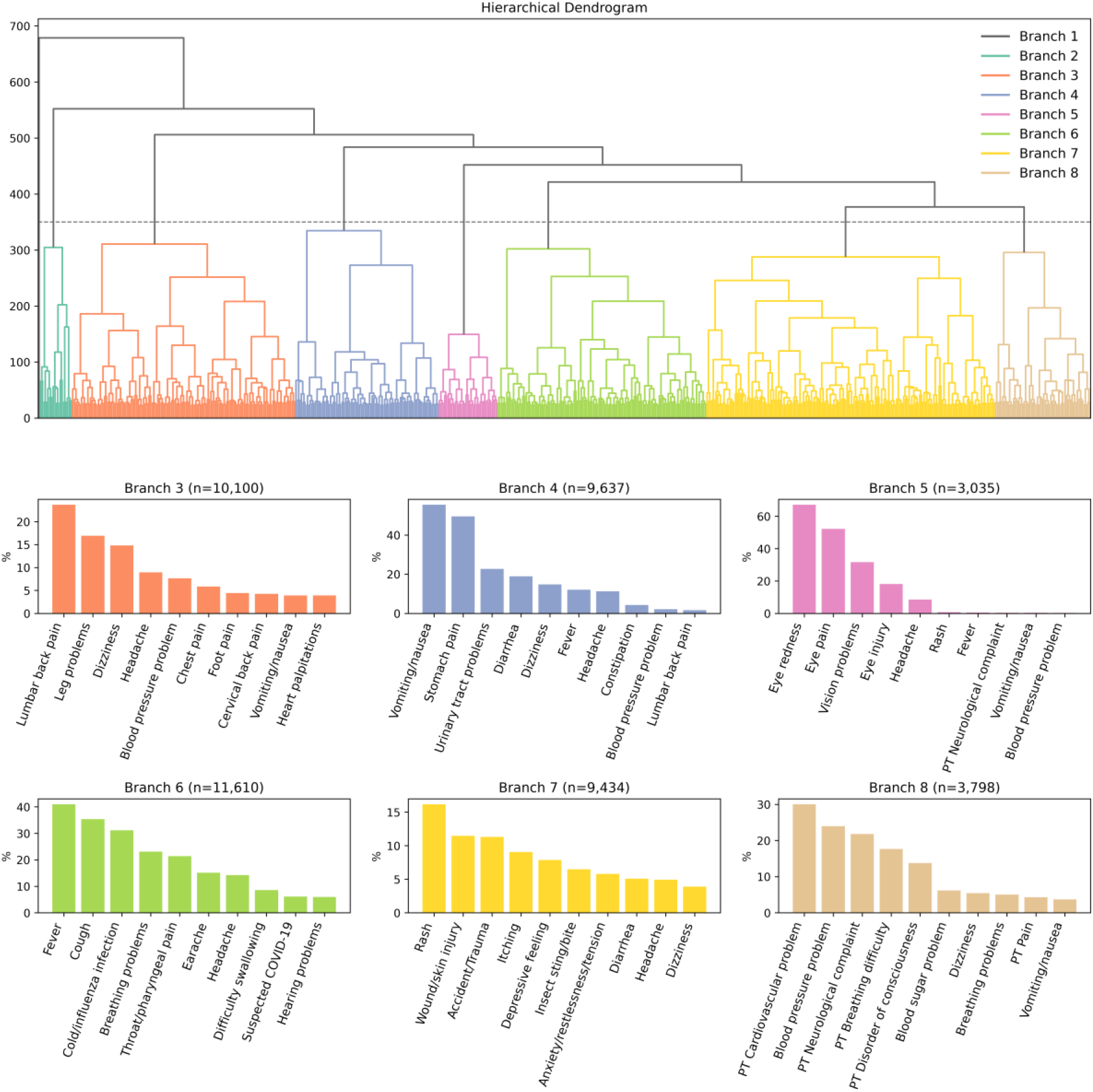
Main branches of the agglomerative clustering of triage assessments and their predominant complaint composition. The hierarchical clustering dendrogram of the 50,000 sampled assessments was cut at height 350 to define the color-coded main branches. For each main branch, the 10 most frequent recorded symptoms and pre-triage (PT) complaints are shown.

These findings suggest that semantic embedding-based clustering produced medically interpretable groupings.

Figure 3 shows the proportion of each TTT category per dendrogram branch. It can be observed that many branches are dominated by a certain TTT category (e.g., “Emergency” and “As soon as possible” assessments in main branch 8 of serious problems identified during pre-triage), even though TTT was not part of the clustering features. This suggests that semantically similar assessments have a tendency to share comparable treatment urgencies.

**Figure 3:**
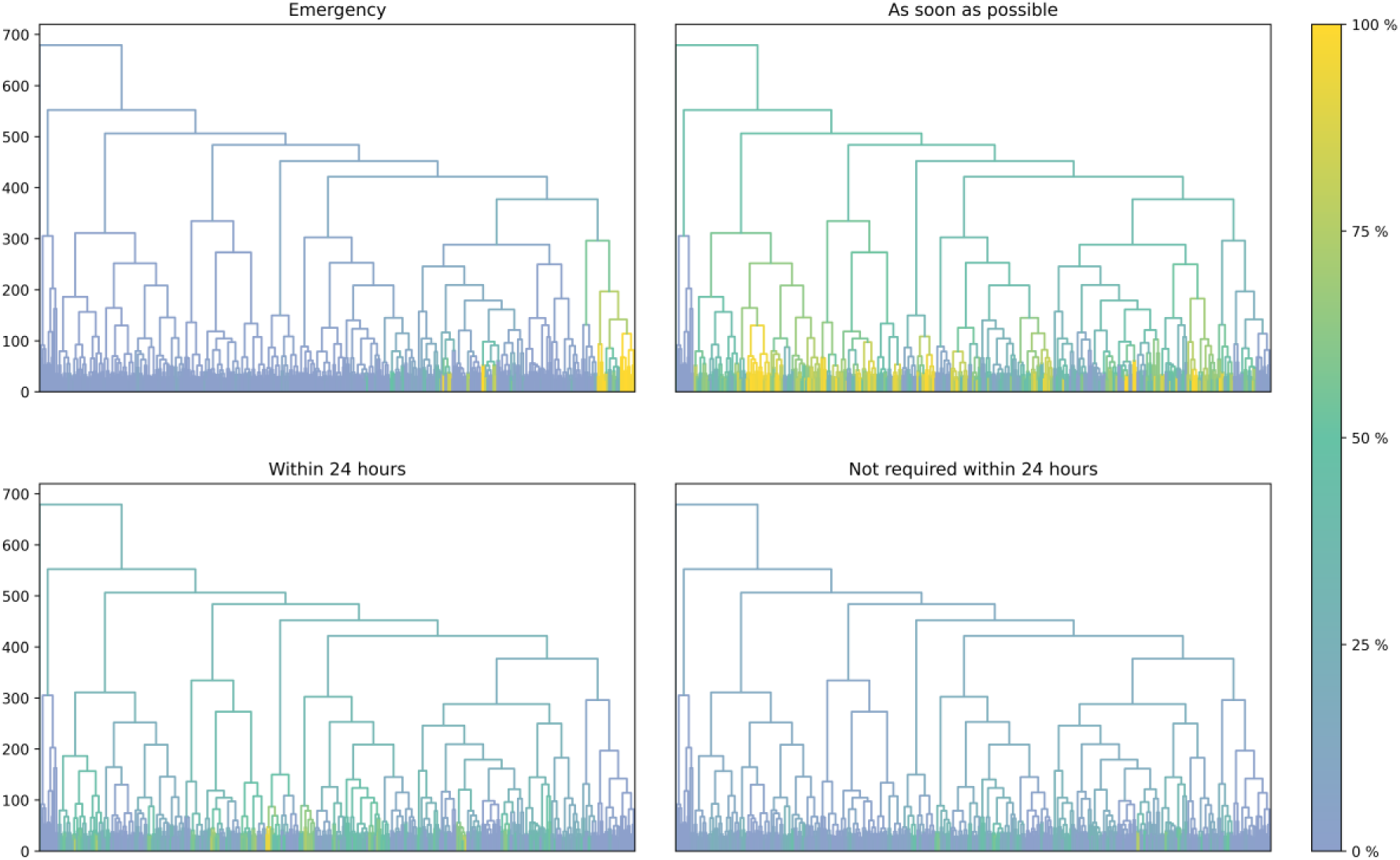
Distribution of TTT categories across dendrogram branches. For each TTT category with explicit urgency assignment, one dendrogram is shown, with branches color-coded according to the proportion of assessments in the respective branch assigned to that TTT category.

#### Cluster Characteristics

The dendrogram was cut at a threshold yielding a TTT homogeneity score of 0.5 (at height 23), resulting in 593 clusters. The assessments were well distributed across clusters; the number of assessments contained in each cluster (mean: 84.3, median: 65, max: 487, min: 6) is shown in Supplementary Figure A.2 in Multimedia Appendix 4. To focus on sufficiently large clusters for representative generation, we considered only clusters containing at least 30 assessments (498 clusters, 48,143 assessments). Increasing this threshold would have reduced sample coverage substantially (see Supplementary Table A.1 in Multimedia Appendix 1). Although emergency cases were less frequent, they are of particular medical relevance.

Therefore, clusters composed entirely of emergency assessments were additionally included, adding 16 clusters comprising 257 assessments. In total, this resulted in 514 clusters, collectively containing 48,400 assessments that represented 96.8% of the original sample. For each of these 514 clusters, we have generated a representative assessment, the development steps of which are presented in the following section.

### Generation and Internal Evaluation of Representative Assessments

For each included cluster, one representative assessment was derived using a two-phase optimization procedure. Phase 1 focused on generating an assessment that captured the central characteristics of the source cluster by minimizing the distance to the cluster centroid. Phase 2 then enriched this assessment with additional symptom-specific details while retaining the symptom set identified in Phase 1. The following analyses evaluate how these representative assessments developed across both optimization phases and how well they reflected their source clusters in terms of their triage urgency.

#### Development Phases

For each of the 514 included clusters, the real assessment with the smallest distance to the cluster centroid was selected as the starting point for representative generation. In Phase 1, the optimization procedure generated a new artificial assessment that was closer to the centroid than the initial assessment in 346 clusters (67.3%). In the remaining 168 clusters (32.7%), the centroid-closest real assessment already represented the best solution. Among all 514 clusters, 458 (89.1%) yielded representative assessments that progressed beyond the pre-triage stage and were assigned at least one symptom profile, making them eligible for the enrichment step in Phase 2. In this second phase, a more comprehensive representative assessment was identified for 289 clusters (56.2% of all clusters), which remained close to the cluster centroid despite added information, while 225 clusters (43.8% of all clusters) retained their Phase 1 representation. Overall, the final set of representative assessments comprised 126 (24.5%) unchanged real assessments corresponding to the initial centroid-closest case and 388 (75.5%) algorithmically generated assessments derived through optimization Phase 1 and/or Phase 2. These findings indicate that the centroid-closest real assessment often provided a useful starting point, but in most clusters the optimization procedure generated a representative assessment that was either more centrally located within the cluster and therefore likely to better reflect cluster-level characteristics, more information-rich, or both.

#### Cluster Centrality and Information Content

To assess cluster centrality and information content, we compared the original cluster assessments, the real centroid-closest assessments, and the generated representative assessments obtained after Phase 1 and 2 (see Figure 4).

**Figure 4:**
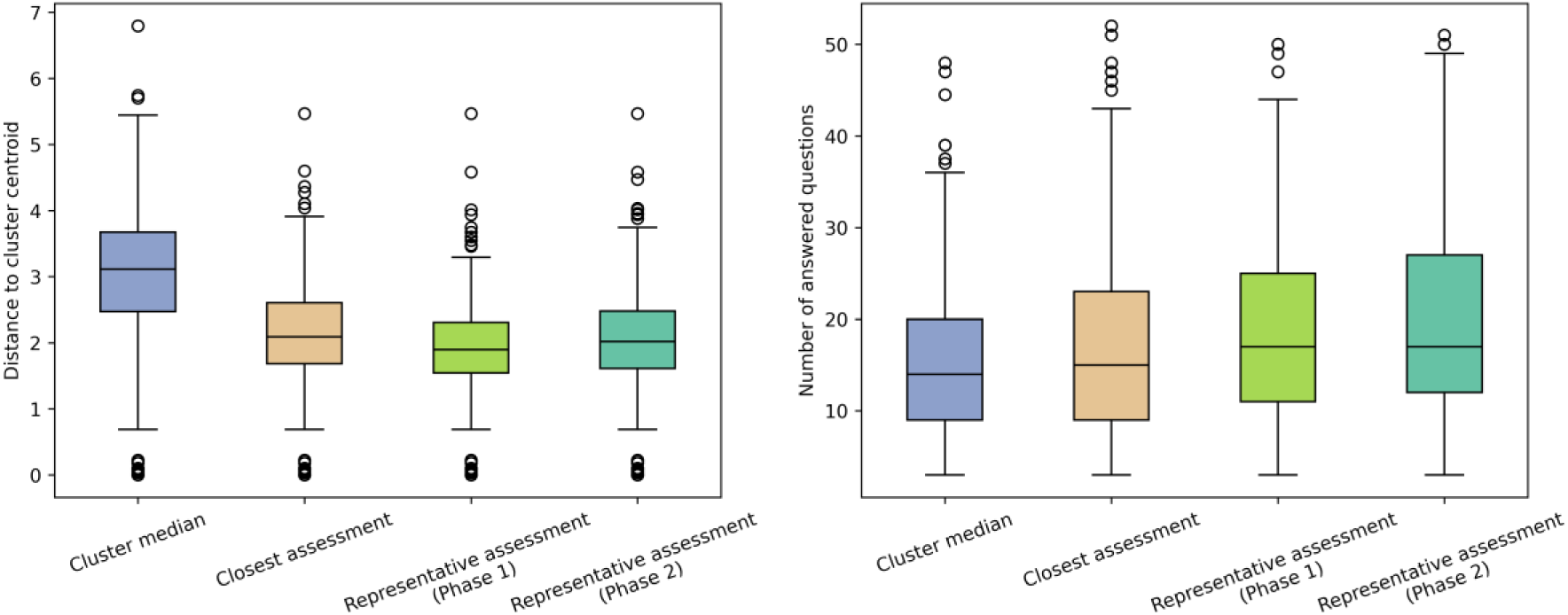
Evolution of representative assessments in terms of distance to the cluster centroid (left) and number of triage questions answered (right). For each cluster, the median value among the original assessments was compared with the centroid-closest real assessment and its development over optimization Phase 1 and Phase 2. Phase 1 reduced the distance to the cluster centroid, whereas Phase 2 increased the number of answered questions while largely preserving cluster centrality.

Across clusters, the median distance of original assessments within a cluster to their centroid was on average 3.1 (median: 3.1, interquartile range (IQR): 2.5-3.7). Selecting the centroid-closest real assessment reduced this distance to an average of 2.1 (median: 2.1, IQR: 1.7-2.6). After Phase 1, this average distance further decreased to 1.9 (median: 1.9, IQR: 1.5-2.3), indicating that the first optimization phase generally improved cluster centrality relative to the closest real assessment available in the cluster. Following Phase 2, the average distance increased slightly to 2.1 (median: 2.0, IQR: 1.6-2.5), showing that the enrichment step resulted in only a modest loss of centrality.

At the same time, information content - quantified as the number of answered triage questions - increased progressively over the two optimization phases. Across clusters, the median number of answered questions among the original assessments was on average of 15.1 (median: 14, IQR: 9-20). The initial centroid-closest real assessment contained on average 16.7 answered questions (median: 15, IQR: 9-23). This number increased to 18.1 (median: 17, IQR: 11-25) after Phase 1 and 19.4 (median: 17, IQR: 12-27) after Phase 2. Compared with the medium number of answered questions among the original assessments within each cluster, the final representative assessments therefore contained on average 4.3 additional answered triage questions.

Taken together, these results indicate that the two-phase optimization procedure achieved the intended trade-off between cluster-centrality and information content.

#### Agreement with Cluster-Level Treatment Urgency

To examine whether the representative assessments preserved clinically relevant urgency patterns of their source cluster, each representative assessment was processed through the triage system, and its TTT recommendation was compared with the TTT recommendations of the originating cluster.

Table 2 summarizes, for each TTT category, the number of clusters in which this TTT represented the majority recommendation (note that the purity of TTT categories varied across clusters), the TTT of the closest initial (real) assessments, and the number of representative assessments from Phase 1 and 2 receiving the same TTT recommendation. The overall distribution of TTT categories among the representative assessments closely resembled the distribution of cluster-level majority TTT categories. The largest absolute deviation was 2.9% for the category “As soon as possible”. When TTT categories were ordinally encoded, the TTT of the final representative assessments (Phase 2) showed a strong positive rank correlation with the cluster’s mean ordinal TTT (Spearman ρ=0.78, p<0.001). Together, these findings suggest that the optimization procedure largely preserved the treatment-urgency patterns of the originating clusters.

**Table 2:** Distribution of cluster-level majority TTT categories and TTT recommendations of the initial and representative assessments.

| TTT | Cluster-level majority (N=514), n (%) | Initial centroid-closest assessment (N=514), n (%) | Representative assessment Phase 1 (N=514), n (%) | Representative assessment Phase 2 (N=514), n (%) |
| --- | --- | --- | --- | --- |
| Emergency | 49 (9.5) | 54 (10.5) | 45 (8.8) | 49 (9.5) |
| As soon as possible | 275 (53.5) | 229 (44.6) | 269 (52.3) | 290 (56.4) |
| Within 24 hours | 135 (26.3) | 151 (29.4) | 140 (27.2) | 121 (23.5) |
| Not required within 24 hours | 41 (8.0) | 65 (12.6) | 39 (7.6) | 39 (7.6) |
| Unclear or N/A | 14 (2.7) | 15 (2.9) | 21 (4.1) | 15 (2.9) |

### Population-Level Representativeness

In this section, we evaluate to what extent the final representative assessments reproduce key characteristics of the 50,000 sampled medical on-call assessments. The 514 clusters used for generating representatives collectively contained 48,400 assessments, corresponding to 96.8% of the sample. To assess potential selection bias due to non-represented clusters, we compare the overall sample distributions with those of the 48,400-assessment subset. Because each representative assessment was derived from a single cluster, the representatives were weighted by the number of assessments contained in their source cluster. For each investigated characteristic, we additionally report the proportion of assessments belonging to clusters in which that characteristic formed the majority value, as an indicator of how strongly it was clustered.

#### Triage Outcomes

We applied the same triage system from which the original assessments were obtained to the generated representative assessments and compared the resulting triage recommendations with those of the full sample and of the assessments contained in the selected clusters. Overall, the distributions of TTT and POC were highly similar between the full sample and the subset of assessments contained in the selected clusters, indicating that exclusion of smaller non-selected clusters introduced only little distortion with respect to these outcomes.

In the cluster-size weighted representative assessments, however, a moderate shift toward higher-urgency categories was observed (see Table 3). Compared with the full sample, the proportion classified as “As soon as possible” increased by 6.6 percentage points (50.5% vs. 43.9%), and “Emergency room” recommendations increased by 7.2 percentage points (33.8% vs. 26.6%). In contrast, the lower-urgency recommendations “Not required within 24 hours” and “Teleconsultation” were underrepresented by 6.3 percentage points (7.7% vs. 14.0%) and 6.5 percentage points (9.8% vs. 16.3%), respectively.

**Table 3:**
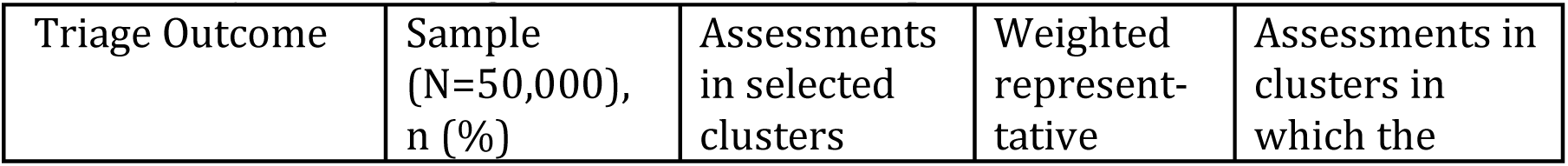
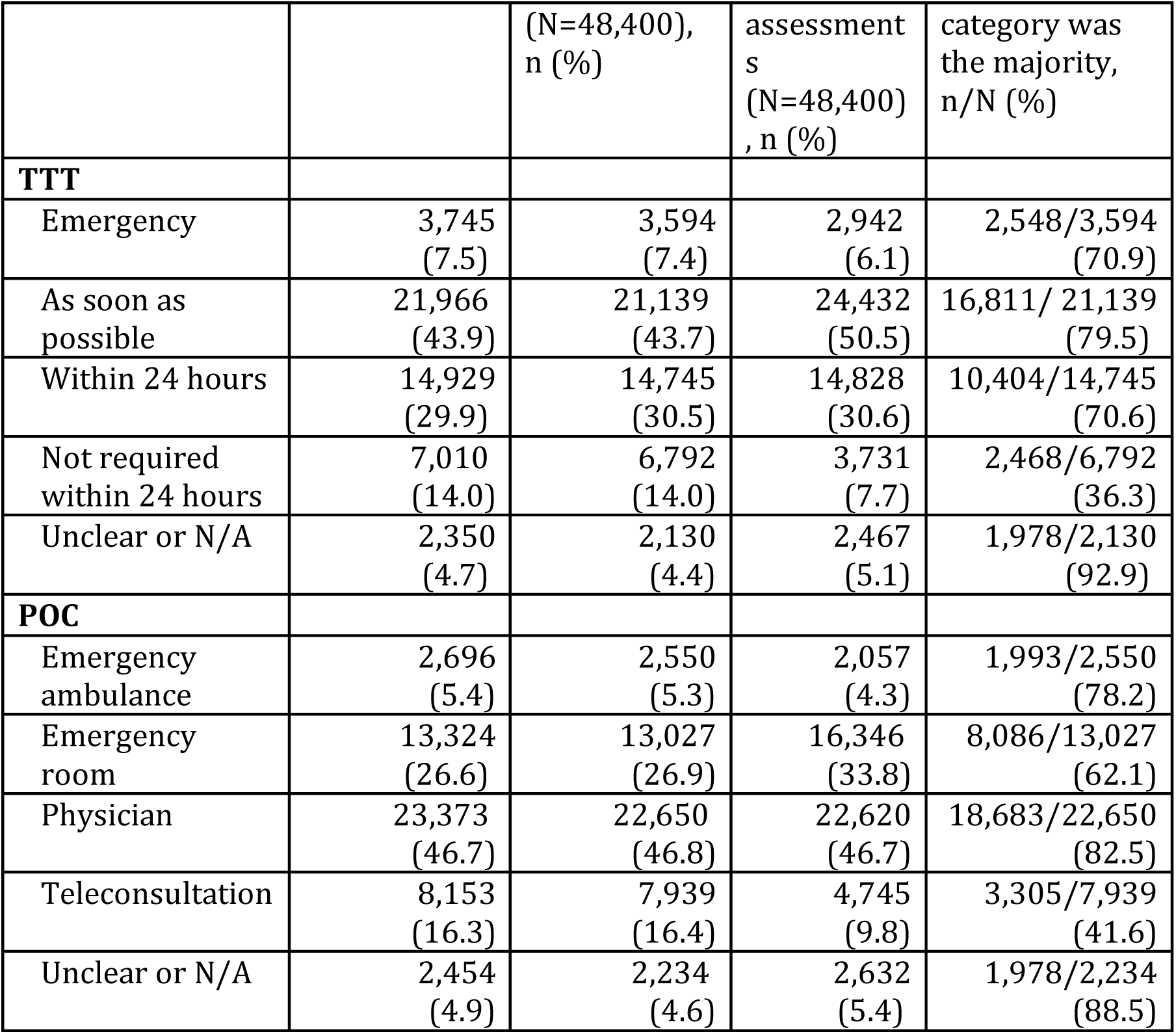
Comparison of triage outcomes of the full 50,000 assessment sample, the assessments within included clusters for representative generation (48,400 assessments), and the weighted set of the final representative assessments.

To help interpret these deviations, we additionally calculated, for each TTT and POC category, the proportion of assessments belonging to clusters in which that category constitutes the majority value. High proportions were observed for “Physician” (82.5%), “As soon as possible” (79.5%) and “Emergency ambulance” (78.2%), indicating that these categories formed relatively coherent cluster majorities. In contrast, only 36.3% of assessments with the recommendation “Not required within 24 hours” belonged to clusters dominated by this category, which may partly explain its underrepresentation in the weighted representative assessments.

#### Demographic Characteristics

The gender distribution of the 48,400 assessments underlying the representative assessments was nearly identical to that of the full 50,000-assessment sample (see Table 4). In contrast, the cluster-size-weighted representative assessments represented 13.5% more female cases than the original sample. Women were overall more frequent in the sample than men (57.4% vs. 42.2%), and 84% of female assessments belonged to clusters in which females formed the majority, compared with 38% for male assessments. As a result, clusters with theoretically gender-neutral symptom patterns tended to contain more female cases, making them more likely to serve as representatives and, when aggregated across clusters, leading to an overall shift toward women in the weighted representative set.

**Table 4:** Comparison of demographic characteristics of the full 50,000 assessment sample, the assessments within included clusters for representative generation (48,400 assessments), and the weighted set of the final representative assessments.

| Demographics | Sample<br>(N=50,000),<br>n (%) | Assessments<br>in selected<br>clusters<br>(N=48,400),<br>n (%) | Weighted<br>representative<br>assessments<br>(N=48,400),<br>n (%) | Assessments in<br>clusters in<br>which the<br>category was<br>the majority,<br>n/N (%) |
| --- | --- | --- | --- | --- |
| **Gender** |  |  |  |  |
| Male | 21,120<br>(42.2) | 20,416<br>(42.2) | 13,607<br>(28.1) | 7,660/20,416<br>(37.6) |
| Female | 28,692<br>(57.4) | 27,800<br>(57.4) | 34,306<br>(70.9) | 23,432/27,800<br>(84.3) |
| N/A | 188<br>(0.4) | 184<br>(0.4) | 487<br>(1.0) | 182/184<br>(98.9) |
| **Age** |  |  |  |  |
| 1-4 Weeks | 82<br>(0.2) | 64<br>(0.1) | 132<br>(0.3) | 0/64<br>(0.0) |
| 5-8 Weeks | 92<br>(0.2) | 65<br>(0.1) | 0<br>(0.0) | 0/65<br>(0.0) |
| 3-12 Months | 873<br>(1.7) | 829<br>(1.7) | 907<br>(1.9) | 357/829<br>(43.1) |
| 1-3 Years | 2,414<br>(4.8) | 2,343<br>(4.8) | 2,590<br>(5.4) | 1,266/2,343<br>(54.0) |
| 4-8 Years | 2,190<br>(4.4) | 2,089<br>(4.3) | 1,466<br>(3.0) | 509/2,089<br>(24.4) |
| 9-13 Years | 1,114<br>(2.2) | 1,042<br>(2.2) | 110<br>(0.2) | 33/1,042<br>(3.2) |
| 14-49 Years | 19,790<br>(39.6) | 19,307<br>(39.9) | 23,057<br>(47.6) | 18,101/19,307<br>(93.8) |
| 50-65 Years | 7,375<br>(14.8) | 7,122<br>(14.7) | 5,532<br>(11.4) | 2,045/7,122<br>(28.7) |
| 66-80 Years | 7,372<br>(14.7) | 7,142<br>(14.8) | 5,325<br>(11.0) | 1,509/7,142<br>(21.1) |
| >80 Years | 8,448<br>(16.9) | 8,164<br>(16.9) | 8,794<br>(18.2) | 4,899/8,164<br>(60.0) |
| N/A | 250<br>(0.5) | 233<br>(0.5) | 487<br>(1.0) | 230/233<br>(98.7) |

The age distribution of the weighted representative assessments was overall similar to that of the sample, although some deviations were observed. The largest deviation occurred in the 14-49-year group, which was overrepresented in the weighted representatives (+8.0%). This likely reflects a similar mechanism as mentioned for the female cases, since this group forms the largest age segment in the sample (39.6%) and 93.8% of assessments in this age group belonged to clusters in which they formed the majority.

#### Symptom Coverage

During the sampling period, the triage system recorded information on 121 distinct symptoms. Out of these, 101 (83.5%) were represented in at least one final generated representative assessment. The remaining, non-covered symptoms occurred in fewer than 0.5% of all sampled assessments. To assess broader symptom coverage, the symptoms were mapped to symptom categories loosely aligned with ICD-10-GM chapters (see Section “Symptom Coverage and Symptom Category Mapping”). For each symptom category, the proportion of assessments that recorded at least one symptom from that category was determined for the total sample (N=50,000) and for the subset of representative clusters (N=48,400) (see “Sample” and “Subsample” in Figure 5). These frequencies were compared with the weighted proportions of representative assessments containing a symptom of the corresponding category (see “Weighted representative assessments” in Figure 5).

**Figure 5:**
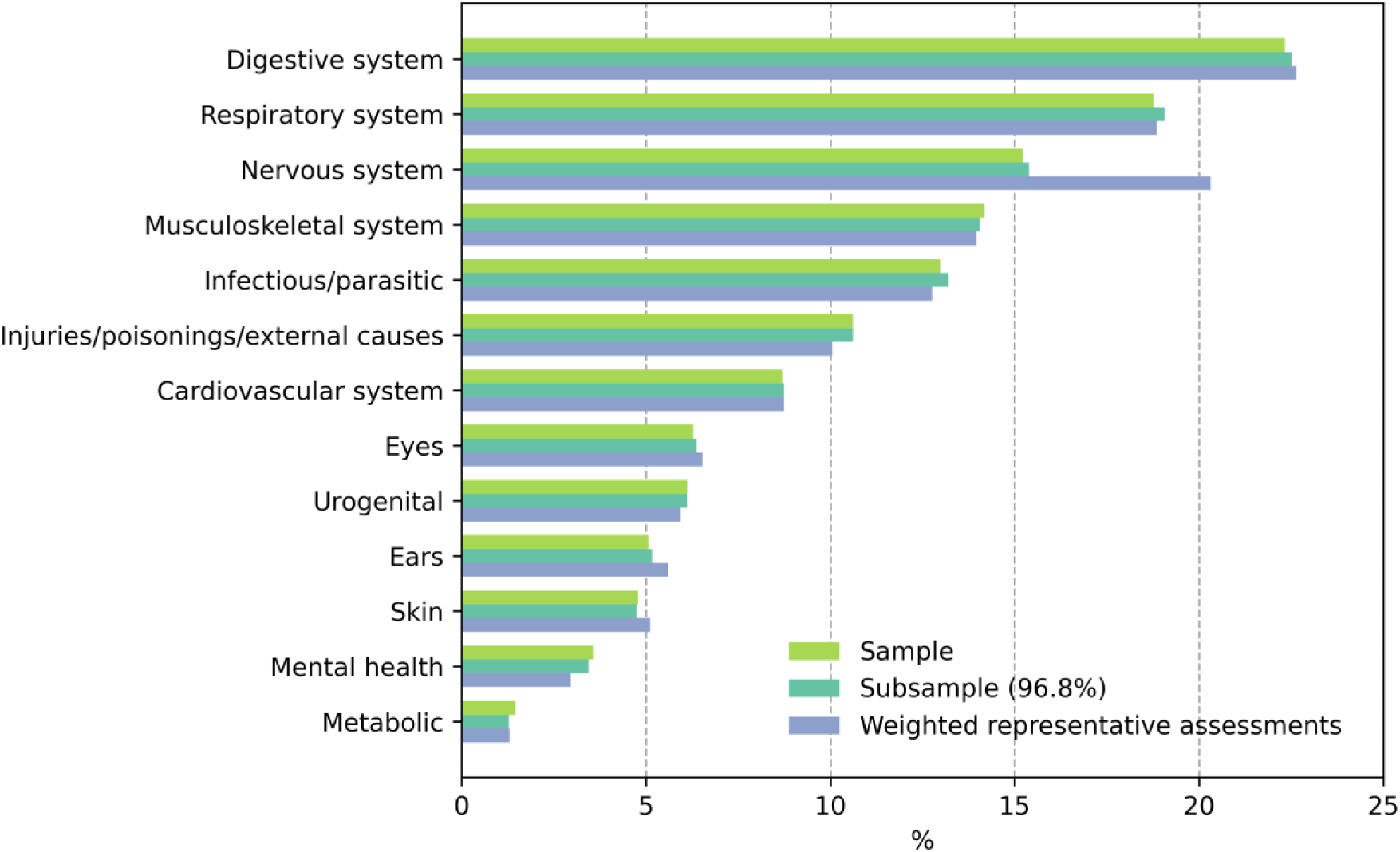
Comparison of the symptom category distributions of the full 50,000 assessment sample, the assessments within included clusters for representative generation (Subsample; 48,400 assessments) and the weighted set of the final representative assessments.

Across most symptom categories, the weighted representative assessments closely approximated the sample frequencies. The largest deviation was observed for the category “Nervous system” (deviation: 5.1%, sample: 15.2%, weighted representatives: 20.3%), mainly driven by the symptom “headache” (+5.3% in the weighted representatives). All remaining symptom categories differed by less than 1%, indicating that the representative assessments closely reproduce the overall symptom distribution of the source data.

#### Sensitivity Analysis of Vignette Weights and Population Coverage

The analyses above evaluated how well the representative assessments reproduced key characteristics of the 50,000-assessment sample when weighted by cluster size. To examine whether these cluster-based, sample-derived vignette weights were preserved at population level, we additionally applied a similarity-based sensitivity analysis to the full 1-year population of approximately 3.2 million assessments.

The resulting similarity-based vignette weights showed a strong positive rank correlation with the original cluster-based weights (Spearman’s ρ=0.77, p<0.001), suggesting that the relative importance of individual vignettes was largely preserved from sample to population.

In the similarity-based analysis, 90.1% of assessments in the 1-year population were matched to at least one vignette from the generated vignette set. As a reference, the original cluster-based approach linked the 514 representative assessments to 48,400 assessments, corresponding to 96.8% of the 50,000-assessment sample.

### Textual Case Vignettes

The representative assessments consisted of sets of question-answer pairs derived from a triage system. These data were transformed into structured medical case vignettes using controlled prompting of an LLM, making them more accessible for healthcare professionals.

The generated vignettes followed the instructed structure, namely 1. Patient information, 2. Medical history and 3. Other medically relevant information. Textbox 1 presents an example vignette (translated from German); additional examples are provided in Supplementary Data A.2 in Multimedia Appendix 5. Information originally recorded by the triage system in interval formats (e.g., age ranges, pain intensity, symptom durations) was successfully converted into specific values (marked by underlining in the text) to simulate realistic patient presentations.

#### Textbox 1

Example of a generated case vignette.

1. Patient: A <u>2-year-old</u> boy.
2. Medical history (anamnesis): Symptom complex: ear discharge, ear pain, fever: For the past <u>2 days</u>, fluid has been coming from the right ear, with no pus or blood visible. In addition, moderate right-sided ear pain (5 out of 10) has been present since today. The parents report that the boy has a fever; the measured body temperature is <u>38.7°C</u>, and the fever began today. Mild chills are reported as an accompanying symptom. The child is more restless than usual. Home remedies or over-the-counter medications have already been used for self-treatment. Chin function is normal; the chin can be moved toward the chest. There is no extreme dryness of the mouth, no marked thirst, and no restriction of urination. Inflammation of the auricle or the area around the ear is unclear. No blisters are visible on the auricle or around the ear.
3. Other medically relevant information: There is no immediately life-threatening situation. No relevant surgery has been performed within the past 14 days. No immune deficiency is known. The young patient has not been abroad recently. He is not currently taking any new medications, and within the past few weeks no new medication has been started and no dosage of existing medications has been increased.

## Discussion

### Principal Findings

This study presents an automated, data-driven framework for condensing a large volume of real triage data from the German medical on-call service (116117) into a human-manageable set of weighted case vignettes linked to clusters covering the vast majority of sampled patient contacts. From a random sample of 50,000 triage assessments, the best clustering performance was achieved using agglomerative clustering with semantic embeddings, resulting in 514 representative clusters that together covered 96.8% of the sample. The resulting dendrogram revealed medically interpretable groupings, such as branches dominated by digestive-tract symptoms, eye complaints, or common cold-like presentations, indicating that semantic embedding-based clustering captured meaningful structure in this heterogeneous triage dataset.

To derive one representative case per cluster, we implemented a two-phase optimization procedure designed to balance proximity to the cluster centroid with information completeness. The resulting representative assessments were, on average, closer to their respective cluster centroids and contained more answered triage questions than the original assessments, indicating improved representativeness and detail. In addition, the TTT recommendations of the representative assessments were strongly correlated with the mean TTT of their source clusters, suggesting that the generated representatives largely preserved clinically relevant triage patterns.

A key contribution of the framework is that each representative assessment remains linked to its source cluster of known size. This makes it possible to assign each vignette an explicit weight within the sample population. When weighted and aggregated across clusters, the set of representative assessments covered all non-rare symptoms captured by the triage system and the full spectrum of treatment-urgency levels. Most demographic, symptom and treatment urgency distributions were approximated reasonably well compared with the original sample, although some systematic deviations remained: most notably an overrepresentation of female cases, individuals aged 14–49 years and the urgency category “As soon as possible”, as well as an underrepresentation of the urgency category “Not required within 24 hours”. These deviations can be explained with the logic of building a single “best” representative case per cluster. Features that cluster well, and therefore dominate certain clusters, are more likely to be included in a representative assessment than features that are widely dispersed across clusters. For instance, only a fraction of assessments with the TTT category “Not required within 24 hours” formed cluster majorities, resulting in fewer representatives of this type than expected from the sample distribution. Similarly, characteristics that are frequent in the source population may become overrepresented, because they more often form cluster majorities and are therefore more likely to be selected in a representative case. An example is gender: females were more common in the sample and therefore more often dominated clusters with gender-neutral symptom patterns, leading to an increased probability to be selected for a representative.

Overall, the generated representatives captured broad presentation diversity of the underlying population, although reducing a large dataset to a single representative case per cluster did not entirely preserve all feature distributions. To assess whether the cluster-based weights were transferable to the full 1-year population, we performed an additional similarity-based sensitivity analysis. Its findings suggested that both broad coverage and the relative importance of individual vignettes were largely preserved at population level.

Finally, the question-answer pairs of the representative assessments were transformed into structured textual case vignettes using a controlled LLM-prompting approach. In this workflow, the LLM was not used to generate case content de novo, but to verbalize information already present in the representative assessments. This step translated triage data into concise, readable case descriptions that may facilitate medical expert review, and further development.

### Comparison with Prior Work

Placed in the broader context of research on clinical vignettes, this study differs from previous work in several aspects. Many prior vignette studies relied on physician-authored vignettes (e.g., [6,9,10]), which can ensure high medical quality but limit the range of clinical presentations represented. Others deliberately focused on predefined disease areas (e.g., [16,24]), thereby narrowing their coverage by design. Approaches that come closer to covering a broad spectrum of medical conditions typically deployed random sampling of real patient cases (e.g., [11–13]). However, only a few explicitly considered population distributions of symptoms or demographics when selecting vignette samples. For example, [12] accounted for the distribution of symptom categories when selecting cases from Reddit posts, representing an exception among largely unweighted sampling strategies.

In contrast, our study used an automated data-driven framework to condense real large-scale triage data into a human-manageable number of representative cases and explicitly evaluated how well key population characteristics were retained. In addition, each generated case remained linked to a quantitative population weight, an aspect that, to our knowledge, has not been addressed in prior vignette studies. This is relevant because explicit weighting may allow vignette-based evaluation results, such as under-triage or over-triage, to be extrapolated to the expected number of affected patients in the underlying population.

With the increasing capabilities of LLMs in recent years, recent studies introduced LLMs into their vignette generation workflow. Some used LLMs primarily to structure or formulate existing medical information (e.g., [25,26]), others employed them to generate vignette content for predefined diseases (e.g., [15,16,18]). Our study belongs to the first category: a representative triage assessment was supplied as factual input, and the LLM was used solely to structure and verbalize this information into medical vignettes. In this sense, our approach illustrates how real data-grounded content and the linguistic capabilities of LLMs can complement each other.

### Limitations

The following limitations should be considered when interpreting the findings of this study.

The generated vignettes are based on assessments from the German medical on-call service (116117) and therefore reflect only patients who accessed this service rather than all patients with acute health concerns in Germany. Use of 116117 may depend on factors such as awareness of the service [27], help-seeking preferences, and access to or ability to use telephone-, web- or on-site triage pathways, which may limit representativeness for some patient groups. In addition, the vignettes are constraint by the information captured by the triage system. This includes predefined symptom lists, their permitted combinations, the level of detail recorded for each symptom, and more general patient information such as pre-existing conditions or medication use. Many responses are also recorded only in broad categories (e.g., age ranges, vital-sign intervals, and duration of symptoms).

The triage system itself is designed to provide rapid, rule-based urgency recommendations rather than capturing complete medical patient histories. As a result, emergency assessments in particular are intentionally short and focused on the core problem to avoid delays in patient treatment. Consequently, the resulting vignettes may lack the narrative richness of real patient encounters.

Furthermore, the optimization algorithm condenses each cluster into a single representative case, thereby reducing intra-cluster variance. Rare or atypical presentations may thus be underrepresented or disappear entirely. In addition, generating a representative assessment close to the cluster centroid may combine characteristics of the cluster that seldom co-occur in real patients. More broadly, semantic similarity in the embedding space does not necessarily imply full medical equivalence.

These limitations were also reflected in a preliminary medical expert review of the generated vignettes. The review identified two main types of issues in some vignettes. First, some cases lacked contextual details that would help clinicians assess them with greater confidence, such as precise durations of symptoms, the circumstances of an accident, or the nature of a wound, as well as overly generalized information originating from the triage system that could represent distinct clinical conditions if specified individually (e.g., “Paralysis/sensory disturbances?”). Second, some vignettes showed logical inconsistencies, such as contradictory timelines between related symptoms or events (e.g., a fall and a head injury) or conflicting information on risk factors. These observations highlight that data-driven representativeness does not automatically guarantee full clinical coherence at the individual vignette level.

The LLM was prompted under strict constraints to avoid hallucinations and to include all available information from the selected question-answer pairs. However, complete control over the generation process cannot be guaranteed.

Finally, because the data used in this study originates from a single national triage system, the extent to which the vignettes generalize to other healthcare settings remains to be evaluated.

### Implications and Future Directions

We condensed thousands of triage assessments into a human-manageable set of 514 representative cases that together characterize almost the entire sample population. Each case vignette is linked to a specific number of underlying assessments, providing a sample-based weight and thereby supporting weighted evaluation frameworks. This kind of linkage between vignettes and underlying assessment frequency has, to our knowledge, not been available in previous vignette datasets.

The resulting large and diverse dataset of vignettes may provide a useful basis for studying differences in triage or diagnostic behavior across various digital decision-support tools, symptom checkers, or LLMs - all of which are increasingly used in medical contexts [1–3] and therefore affect a growing number of patients. The large number of vignettes may enable more stratified comparison than smaller vignette sets. Applying the vignettes to other digital triage systems may highlight limitations in our present narratives encapsulated in the question-answer sequences as generated by the on-call medical service in Germany. Enriching the present data basis may increase the value of the resulting set of vignettes.

Building on the findings of the initial medical review, planned next steps include iterative physician reviews of all vignettes to improve their clinical coherence and validity, followed by physician triage of the finalized set of vignettes. We aim to release the curated vignette set as a public benchmark dataset and to enable submission of results from triage tools for comparison with medical expert consensus. These steps will not only enable systematic comparison between tools and human judgment but also allow assessment of potential risks such as under- or over-triage. Beyond benchmarking, the vignettes may serve as a valuable resource for physician education and training and could contribute to the further development of safe and reliable triage systems.

### Conclusions

This study presents a data-driven framework for generating a human-manageable set of population-linked case vignettes from nationwide triage data. Applied to a random sample of 50,000 triage assessments, the framework produced 514 weighted representative cases that covered most sampled presentations and approximated key distributions of demographics, symptom categories, and treatment urgency. The generated vignettes contribute to the diversity of existing vignette resources and may support more population-aware benchmarking of digital decision-support tools. Before publishing the vignette set, expert refinement is planned to strengthen medical coherence and quality.

## Supporting information

Multimedia Appendix 1

Multimedia Appendix 2

Multimedia Appendix 3

Multimedia Appendix 4

Multimedia Appendix 5

## Acknowledgements

We declare the use of GPT-5.4 (OpenAI) for language editing of the manuscript.

## Data Availability

The triage data used in this study are not publicly available because they contain patient health information and are subject to legal and data protection restrictions. The derived vignette dataset is not publicly available at the time of publication because it is still undergoing medical expert review and refinement to improve clinical coherence and quality. Public release of a curated vignette dataset is planned for a later stage, in accordance with applicable legal, governance, and data protection requirements.

## Authors’ Contributions

Conceptualization: AS, LK, ES, JS

Data curation: AS

Formal analysis: AS

Funding acquisition: LK

Investigation: AS

Methodology: AS, LK, ES, JS

Project administration: LK

Resources: LK

Software: AS

Supervision: LK, ES

Validation: AS

Visualization: AS

Writing – original draft: AS

Writing – review & editing: AS, ES, LK, JS

## Abbreviations

ICD-10-GM: 10th revision of the International Classification of Diseases – German Modification
IQR: interquartile range
LLM: large language model
PCA: principal component analysis
POC: point of care
PT: pre-triage
SD: standard deviation
TTT: time to treat

