## Supplementary material for "A Data-Driven Framework for Generating Population-Linked Case Vignettes from Nationwide Triage Data": Multimedia Appendix 1

Comparison of different agglomerative clustering configurations and the sensitivity to the minimum cluster size and the trade-off parameter λ for representative assessment generation.

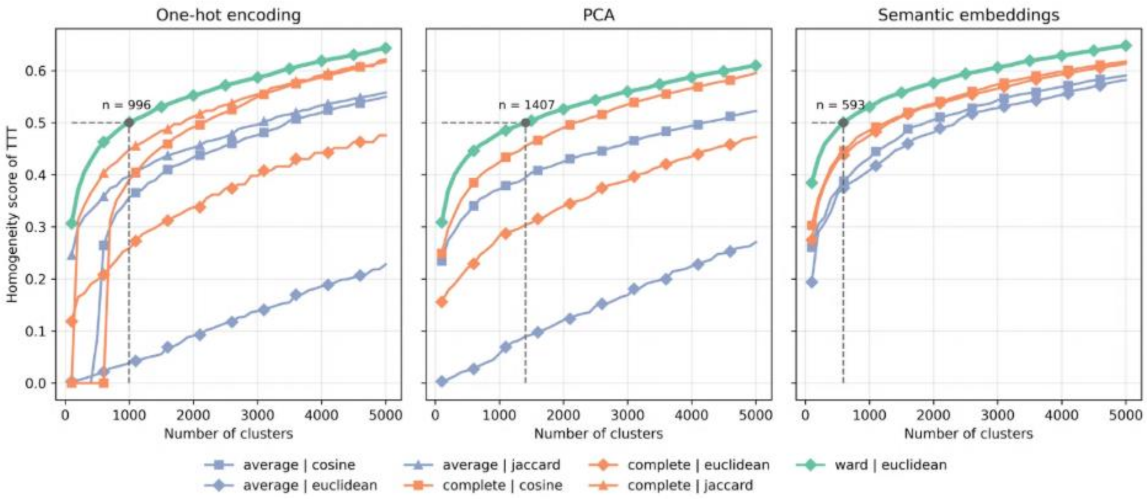


Supplementary Figure A.1: Homogeneity of TTT by the number of clusters for different agglomerative clustering configurations. Question-answer pairs from triage assessments were represented as one-hot encoded vectors (3,709 dimensions), one-hot encoded vectors followed by PCA retaining at least 90% variance (473 dimensions), or semantic embeddings generated with the Sentence Transformer Model "German Semantic STS V2" (1,024 dimensions). Eligible combinations of the linkage methods ("average", "complete", "ward") and distance metrics ("cosine", "jaccard", "euclidean") were evaluated. The Sentence Transformer embeddings combined with Ward linkage and Euclidean distance achieved a TTT homogeneity score of 0.5 with the fewest clusters (n=593) and were therefore selected for subsequent analysis.

Supplementary Table A.1: Sensitivity of cluster inclusion and covered assessments to the minimum cluster-size threshold. For the selected clustering configuration, “German Semantic STS V2” with Ward linkage and Euclidean distance, the number and proportion of clusters retained and sampled assessments covered are shown for different minimum cluster-size thresholds.

| Minimum cluster size | Retained clusters (N=593), n (%) | Covered assessments (N=50,000), n (%) |
|---|---|---|
| 30 | 498 (84.0) | 48,143 (96.3) |
| 40 | 433 (73.0) | 45,892 (91.8) |
| 50 | 377 (63.6) | 43,431 (86.9) |

Supplementary Table A.2: Sensitivity of the trade-off parameter λ used to balance cluster centrality with symptom-detail in the representative assessment generation Phase 2. A value of λ=0.5 weights cluster-proximity and answered questions equally, while λ=0.25 favors more answered triage questions.

| λ | Representative assessments of Phase1 eligible for Phase 2 enrichment, n (%) | Representative assessments with detail enrichment after Phase 2, n (%) | Distance to the cluster centroid across all final representatives (N=514), mean, median (IQR) | Answered triage questions across all final representatives (N=514), mean, median (IQR) |
|---|---|---|---|---|
| 0.25 | 458 (89.1) | 289 (56.2) | 2.1, 2.0 (1.6-2.5) | 19.4, 17.0 (12.0-27.0) |
| 0.50 | 458 (89.1) | 212 (41.2) | 2.0, 1.9 (1.5-2.3) | 18.7, 17.0 (11.0-26.0) |
