## Supplementary material for "A Data-Driven Framework for Generating Population-Linked Case Vignettes from Nationwide Triage Data": Multimedia Appendix 2

Mapping of triage symptoms to symptom categories.

Supplementary Table A.3: Mapping of 121 symptoms recorded by the triage system (translated from German) to 13 broader symptom categories. Three symptoms could not be assigned to a specific symptom category due to their general nature and are therefore listed as "N/A" in the symptom category column.

| Symptom category | Symptom |
|---|---|
| Cardiovascular system | Lymph node problems |
| Cardiovascular system | Blood pressure problem |
| Cardiovascular system | Chest pain |
| Cardiovascular system | Heart palpitations |
| Cardiovascular system | Heart rhythm disorders |
| Cardiovascular system | Fainting |
| Digestive system | Jaundice |
| Digestive system | Rectal bleeding |
| Digestive system | Anal pain |
| Digestive system | Stomach pain |
| Digestive system | Diarrhea |
| Digestive system | Vomiting/nausea |
| Digestive system | Swallowed a foreign object |
| Digestive system | Mouth, tongue, or lip problems |
| Digestive system | Black stool/Blood in the stool |
| Digestive system | Lower abdominal pain |
| Digestive system | Indigestion |
| Digestive system | Constipation |
| Digestive system | Tooth/gum problems |
| Digestive system | Groin pain |
| Ears | Hearing problems |
| Ears | Foreign object in the ear |
| Ears | Ear discharge |
| Ears | Earache |
| Ears | Ringing in the ears / Tinnitus |
| Eyes | Eye redness |
| Eyes | Eye pain |
| Eyes | Vision problems |
| Infectious/parasitic | Fever |
| Infectious/parasitic | Pubic lice |
| Infectious/parasitic | Head lice |
| Injuries/poisonings/external causes | Vaccine reaction |
| Injuries/poisonings/external causes | Arm injury (due to an accident) |

| | |
|---|---|
| Injuries/poisonings/external causes | Eye injury |
| Injuries/poisonings/external causes | Abdominal injury |
| Injuries/poisonings/external causes | Leg injury (due to an accident) |
| Injuries/poisonings/external causes | Chest injury |
| Injuries/poisonings/external causes | Inhalation foreign object/toxic substance |
| Injuries/poisonings/external causes | Elbow injury (due to an accident) |
| Injuries/poisonings/external causes | Finger injury (due to an accident) |
| Injuries/poisonings/external causes | Foot injury (due to an accident) |
| Injuries/poisonings/external causes | Facial injury |
| Injuries/poisonings/external causes | Plaster-related issues |
| Injuries/poisonings/external causes | Wrist injury (due to an accident) |
| Injuries/poisonings/external causes | Hand injury (due to an accident) |
| Injuries/poisonings/external causes | Hip injury (due to an accident) |
| Injuries/poisonings/external causes | Insect sting/bite |
| Injuries/poisonings/external causes | Knee injury (due to an accident) |
| Injuries/poisonings/external causes | Head injury |
| Injuries/poisonings/external causes | Nose injury |
| Injuries/poisonings/external causes | Back injury |
| Injuries/poisonings/external causes | Shoulder injury (due to an accident) |
| Injuries/poisonings/external causes | Sunstroke / Heat-related illnesses |
| Injuries/poisonings/external causes | Ankle injury (due to an accident) |
| Injuries/poisonings/external causes | Electrical accident / electric shock |
| Injuries/poisonings/external causes | Accident/Trauma |
| Injuries/poisonings/external causes | Burn/Scald |
| Injuries/poisonings/external causes | Poisoning |
| Injuries/poisonings/external causes | Wound/skin injury |
| Injuries/poisonings/external causes | Toe injury (due to an accident) |
| Mental health | Anxiety/restlessness/tension |
| Mental health | Depressive feeling |
| Mental health | Panic attack |
| Mental health | Sleep disorders |
| Mental health | Suicidal thoughts/Suicidal tendencies |
| Mental health | Strange thoughts |
| Mental health | Strange behavior |
| Metabolic | Loss of appetite |
| Metabolic | Blood sugar problem |
| Metabolic | Change in weight |
| Musculoskeletal system | Leg problems |
| Musculoskeletal system | Elbow pain |
| Musculoskeletal system | Finger problems |
| Musculoskeletal system | Foot pain |

| | |
|---|---|
| Musculoskeletal system | Joint pain |
| Musculoskeletal system | Hand problems |
| Musculoskeletal system | Wrist pain |
| Musculoskeletal system | Hip pain |
| Musculoskeletal system | Knee problems |
| Musculoskeletal system | Back/Thoracic Spine Pain |
| Musculoskeletal system | Lumbar back pain |
| Musculoskeletal system | Cervical back pain |
| Musculoskeletal system | Shoulder pain |
| Musculoskeletal system | Ankle pain |
| Musculoskeletal system | Toe problems |
| Musculoskeletal system | Arm pain |
| Nervous system | Headache |
| Nervous system | Neurological seizure |
| Nervous system | Dizziness |
| Nervous system | Speech disorder |
| Nervous system | Facial pain |
| Nervous system | Muscle weakness |
| Respiratory system | Asthma symptoms |
| Respiratory system | Breathing problems |
| Respiratory system | Cold/influenza infection |
| Respiratory system | Suspected COVID-19 |
| Respiratory system | Throat/pharyngeal pain |
| Respiratory system | Cough |
| Respiratory system | Nosebleed |
| Respiratory system | Foreign object in the nose |
| Respiratory system | Difficulty swallowing |
| Urogenital | Penile discharge |
| Urogenital | Vaginal discharge |
| Urogenital | Gynecological symptoms |
| Urogenital | Genital discomfort in men |
| Urogenital | Blood in semen |
| Urogenital | Urinary tract problems |
| Urogenital | Pain in the testicles or scrotum |
| Urogenital | Vaginal bleeding |
| Urogenital | Foreskin problem |
| Urogenital | Breast problem |
| Skin | Abscess |
| Skin | Allergic symptoms |
| Skin | Rash |
| Skin | Dark spot on the skin |

| | |
|---|---|
| Skin | Itching |
| Skin | Lump/Swelling |
| Skin | Sunburn |
| N/A | Fatigue |
| N/A | Feeling unwell |
| N/A | Crying baby/toddler |
