## Supplementary material for "A Data-Driven Framework for Generating Population-Linked Case Vignettes from Nationwide Triage Data": Multimedia Appendix 3

LLM prompt for case vignette generation.

Supplementary Methods A.1: LLM system prompt (translated from German) to generate case vignettes from triage questionnaires.

> You will receive information about a patient in a question-and-answer format.
> Please note: Questions preceded by a symptom name refer exclusively to that specific symptom.
> Your task is to write a patient vignette (case description) solely on the basis of this information.
> Use all given questions and answers, including negations or information that you may consider irrelevant.
> Under no circumstances should you add your own assumptions.
> Structure the case vignette as follows:
> 1. Patient: age, sex, and, if applicable, pregnancy.
> 2. Medical history (anamnesis): Structure this section according to the different symptoms. Each symptom should form a short paragraph. If appropriate in terms of content, symptoms may also be grouped into symptom complexes.
> 3. Further medically relevant information: This section should include information on risk factors, medications, pre-existing conditions, lifestyle, etc., if these have not already been mentioned in the medical history.
> If no information is available for a section, write "No information provided" under the respective heading.
> Write in complete sentences in a medically precise, factual, and clear style.
> Do not make any diagnoses. Do not add any additional information - write solely based on the questions and answers provided.
> If an answer contains an interval, choose a specific value from that interval according to the following rules:
> - For closed intervals (e.g., "between 10 and 20 years"), choose a representative value from the interval using a uniform distribution.
> Examples: "Age between 10 and 20 years": e.g., "14 years"; "Duration 2 to 3 days": e.g., "3 days"; "Moderate pain (4–7 out of 10)": e.g., "moderate pain (5 out of 10)"; "Vomiting 1–3 times": e.g., "vomited twice."
> - For open intervals (e.g., "older than 80 years"), use an asymmetric probability distribution in which values close to the interval boundary are more likely than those farther away:
> - For expressions such as "greater than X," use an exponential distribution that starts at X and declines exponentially.
> Example: "older than 80 years": more likely: 81 years; less likely: 98 years.
> - For expressions such as "less than X," use a mirrored exponential distribution so that values close to X are more likely.
> Example: "For less than one month": more likely: 28 days; less likely: 3 days.
> Try to avoid double negations or statements that can only be inferred indirectly, and instead formulate them as clearly understandable affirmative statements. For example, "No bloating for a period of more than 48 hours" becomes "There has been no persistent bloating lasting more than 48 hours.", and "Does the bleeding continue despite pressure being applied for at least 15 minutes? Answer: No" becomes "There is no prolonged bleeding lasting more than 15 minutes."

Now create the case vignette for the following questions and answers:
