## Supplementary material for "A Data-Driven Framework for Generating Population-Linked Case Vignettes from Nationwide Triage Data": Multimedia Appendix 4

Triage assessments per cluster.

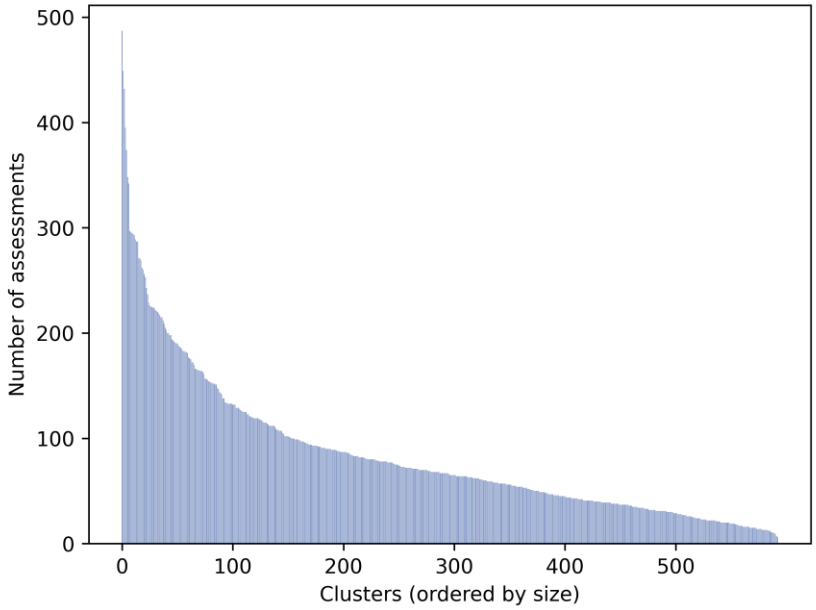


Supplementary Figure A.2: Number of triage assessments per cluster. Clusters (n=593) were obtained by cutting the dendrogram at a threshold yielding a TTT homogeneity score of 0.5.
