## Supplementary material for "A Data-Driven Framework for Generating Population-Linked Case Vignettes from Nationwide Triage Data": Multimedia Appendix 5

Examples of case vignettes.

Supplementary Data A.2: Selection of five case vignettes (translated from German).

1. Patient: The patient is a 61-year-old man.
2. Medical history (anamnesis): Leg complaints: The patient reports bilateral leg complaints that have been present for 4 days. The complaints are described as severe (8 out of 10). There is newly developed swelling of both legs. There are no indications of inflammation such as warmth or redness. Signs of circulatory disturbance were also denied. Weight-bearing on the legs is currently not possible.
3. Further medically relevant information: There are no known risk factors for the development of blood clots (thrombosis or embolism) in the leg, arm, or lung. No further information is available.

---

1. Patient: The patient is an 81-year-old man.
2. Medical history (anamnesis): Abdominal pain and digestive disorder: The patient complains since today of severe, cramping or colicky abdominal pain that is poorly localized and radiates to the back. In addition, he reports a digestive disorder. The pain in the context of the digestive disorder is localized in the middle upper abdomen and is perceived as painful. Furthermore, the patient suffers from heartburn or acid regurgitation. There is flatulence, but not prolonged (>48 hours), and intestinal gas is still being passed. A recently performed abdominal operation as well as a stomach or intestinal endoscopy have not taken place in the past few days. The patient reports no ingestion of spoiled food.
3. Further medically relevant information: The patient has so far undertaken self-treatment measures using home remedies or over-the-counter medications. No relevant previous illnesses, specific risk factors, or known underlying diseases are reported.

---

1. Patient: The patient is a 63-year-old woman.
2. Medical history (anamnesis): Symptom of eye pain: Moderate eye pain (5 out of 10) has been present for 3 days. A specific eye was not mentioned. A small, painful, reddened, pus-filled nodule at the eyelid margin (stye) is present. The eyelids are reddened and swollen, but the eye can be opened. There are no stuck-together eyes or visible pus on the eyes. There are no indications of a foreign body in the eye, prior eye surgeries within the last 14 days, known eye diseases, or recent intensive light exposure (for example at the sea, in snow, due to electric welding or laser). No measures of self-treatment, such as home remedies or over-the-counter medications, have been taken.
3. Further medically relevant information: The patient wears contact lenses. There is no acutely life-threatening condition.

---

1. Patient: Female, 81 years old
2. Medical history (anamnesis): Fall event: The patient fell today. The fall occurred from a height lower than her own body height. There was no loss of consciousness. After the fall, walking and standing remain possible. There are no

indications of movement disorders or skin sensory disturbances such as numbness or tingling.

3. Other medically relevant information: The patient is taking anticoagulant medication or has a known blood coagulation disorder. There is no immediately life-threatening situation.

1. Patient: The patient is a 68-year-old female.
2. Medical history (anamnesis): Cardiovascular complaints: The patient reports newly occurred cardiovascular symptoms. Specifically, she presents with a pale, sallow facial color and is cold-sweaty. She also reports recently developed severe cardiovascular complaints. The patient is breathing adequately and shows a normal response to verbal stimuli.
3. Other medically relevant information: No information provided.
